# Assessing the Limitations of Large Language Models in Clinical Practice Guideline-concordant Treatment Decision-making on Real-world Data

**DOI:** 10.1101/2024.11.20.24313385

**Authors:** Tobias Roeschl, Marie Hoffmann, Djawid Hashemi, Felix Rarreck, Nils Hinrichs, Tobias D. Trippel, Matthias I Gröschel, Axel Unbehaun, Christoph Klein, Jörg Kempfert, Henryk Dreger, Benjamin O’Brien, Gerhard Hindricks, Felix Balzer, Volkmar Falk, Alexander Meyer

## Abstract

**Aims:** Large Language Models (LLMs) have shown promise in therapeutic decision-making comparable to medical experts, but these studies have used highly curated patient data. The aim of this study was to determine whether LLMs can make guideline-concordant treatment decisions based on patient data as it typically presents in clinical practice.

**Methods and Results:** We conducted a retrospective study of 80 patients with severe aortic stenosis who were scheduled for either surgical (SAVR, n=24) or transcatheter aortic valve replacement (TAVR, n=56) by our institutional heart team in 2022. Various LLMs (BioGPT, GPT-3.5, GPT-4, GPT-4 Turbo, GPT-4o, Llama-2, Mistral, PaLM 2, and DeepSeek-R1) were queried using either anonymized original medical reports or manually generated case summaries to determine the most guideline-concordant treatment. Agreement with the Heart Team was measured using Cohen’s kappa coefficients, reliability using intraclass correlation coefficients (ICCs), and fairness using frequency bias indices (FBIs) with FBIs >1 indicating bias towards TAVR. When presented with original medical reports, LLMs showed poor performance (kappa: −0.47–0.22, ICC: 0.0–1.0, FBI: 0.95–1.51). The LLMs’ performance improved substantially when case summaries were used as input and additional guideline knowledge was added to the prompt (kappa: −0.02–0.63, ICC: 0.01–1.0, FBI: 0.46–1.23). Qualitative analysis revealed instances of hallucinations in all LLMs tested.

**Conclusion:** Even advanced LLMs require extensively curated input for informed treatment decisions. Unreliable responses, bias and hallucinations pose significant health risks and highlight the need for caution in applying LLMs to real-world clinical decision-making.

## Introduction

Large Language Models (LLMs) have recently demonstrated their impressive capabilities in medicine, exemplified by passing medical board exams^1^, making correct diagnoses in complex clinical cases^2^ and excelling in physician-patient communication.^3^

Most recently, the use of LLMs in therapeutic decision-making has been trialed. Several studies have shown that LLMs can make treatment decisions for patients with oncological and cardiovascular diseases that are in substantial agreement with the respective treatment decisions made by clinical experts in tumor boards^4–7^ and Heart Teams (HTs)^8^. However, a common feature of these studies was that the LLMs did not make treatment decisions based on real-world patient data in its original format (e.g., discharge letters, imaging reports, etc.) but based on pre-processed data.

In clinical practice, relevant patient data such as patient characteristics, comorbidities, tumor stages and imaging results are typically available in free-text format, either as medical text reports or as text entries in the electronic health record – a format that is likely to persist in the near future. In the aforementioned studies, however, decision-relevant patient data were extracted from the original medical reports by the investigators in a pre-processing step before being provided to the LLMs as input in a concise and high-quality form. However, it is still unknown to what extent LLMs can make treatment decisions based on the original medical data, a scenario that could lead to a significant reduction in physician workload and potentially increase guideline adherence and thus improve patient care.

In this study, we investigate the impact of data representation, i.e. using original medical reports versus case summaries on the performance of LLMs in therapeutic decision-making.

As our study population, we selected patients with severe aortic stenosis (AS). This cohort was chosen because the parameters relevant to decision-making are readily quantifiable, the potential for resource optimization is substantial, and the prevalence of the condition is increasing. If left untreated, AS is associated with high morbidity and mortality.^9^ Treatment modalities for severe AS include surgical aortic valve replacement (SAVR), transcatheter aortic valve replacement (TAVR) and, to a lesser extent, medical therapy. The choice of the optimal treatment modality depends on several clinical variables, including patient age, estimated surgical risk, comorbidities and anatomical factors, as specified in the 2021 guidelines of the European Society of Cardiology (ESC) and the European Association for Cardio-Thoracic Surgery (EACTS) guidelines for the management of valvular heart disease.^10^ The 2021 ESC/EACTS Guidelines strongly endorse an active, collaborative consultation with the multidisciplinary Heart Team (HT). HTs are comprised of cardiologists, cardiac surgeons, cardiac imaging specialists and cardiac anesthesiologists. In HT meetings, these experts review a patient’s condition based on patient data laboriously extracted from medical reports before arriving at a treatment decision using a guideline-based approach.

## Methods

We presented patient data to an LLM to obtain a treatment decision of either SAVR or TAVR. We assessed the degree of agreement between the treatment decisions provided by the LLM and the treatment decisions provided by the HT. Furthermore, we assessed decidability, reliability and fairness of the LLMs. Finally, we compared the performance of seven state-of-the-art LLMs to the performance of a simple non-LLM reference model. In an ablative manner, we studied the effect of using case summaries instead of the original medical reports and adding guideline knowledge to the prompt, respectively, resulting in four distinct experiments (Figure 1).

**Figure 1:**
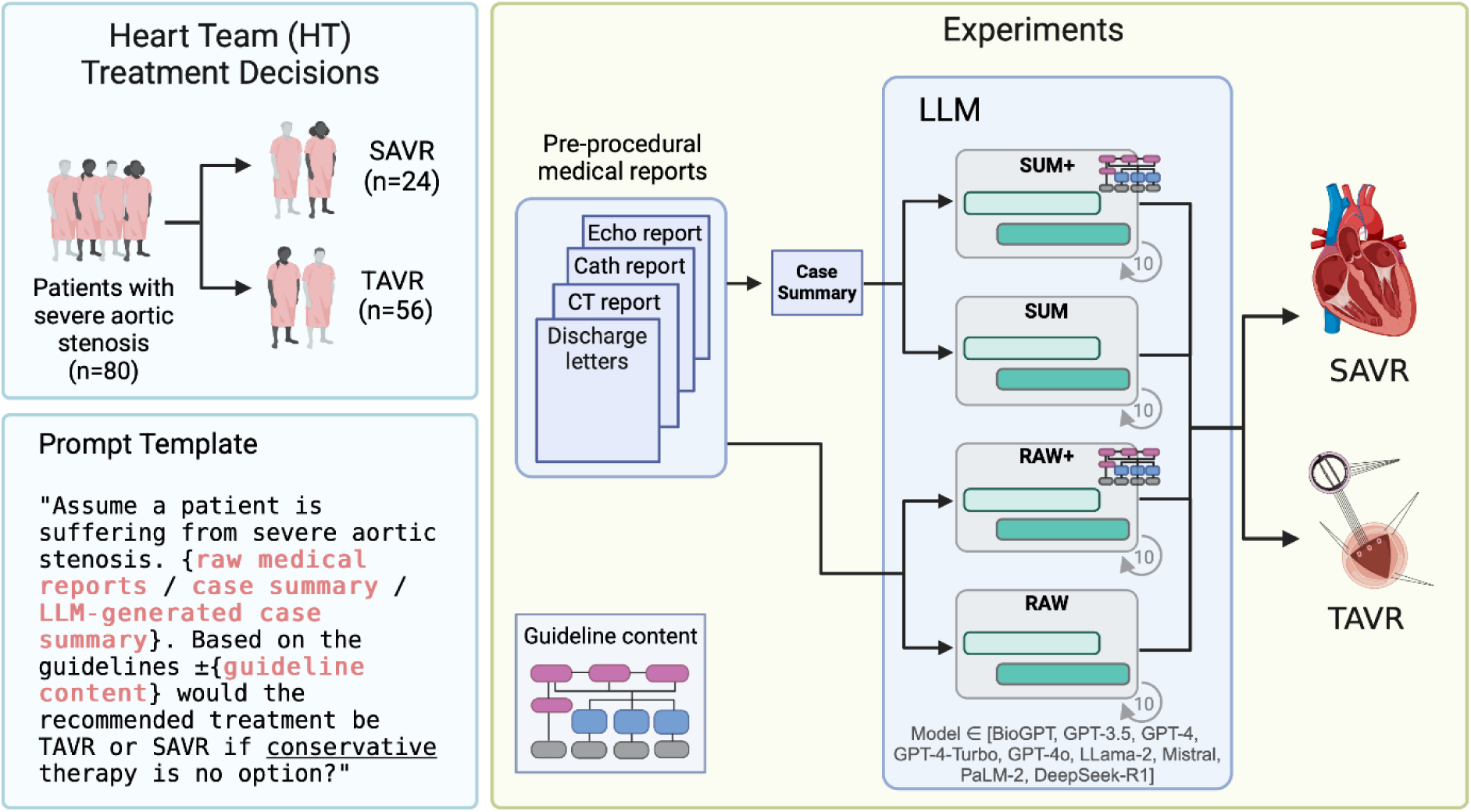
Experimental design. We presented the clinical data of 80 patients suffering from severe aortic stenosis (AS) to a Large Language Model (LLM) to receive a treatment decision for either surgical aortic valve replacement (SAVR) or transcatheter aortic valve replacement (TAVR), repeating each query 10 times. To investigate whether injecting guideline knowledge (RAW+) into the prompt and/or using case summaries (SUM, SUM+) instead of the original medical reports (RAW) improves LLM performance, we conducted a total of four experiments. Case summaries included only decision-relevant patient data and were manually created by physicians.

### Study population

This study included patients treated at a heart center. We screened all patients with severe degenerative AS who were scheduled for a HT meeting in our hospital information system at one campus of our center in 2022. We identified 80 patients with sufficiently digitized documentation. As part of a quaternary care center, our institutional HT receives preselected patients scheduled for invasive AS treatment. Therefore, the number of patients recommended for conservative treatment at our institution is negligible. As a result, we decided to limit the possible therapeutic options for this study to SAVR and TAVR, excluding conservative therapy. This study was approved by the research ethics committee of Charité – Universitätsmedizin Berlin (EA1/146/23).

### Data collection

Medical reports were available as Portable Document Format (PDF) files in our hospital information system. For each patient, we included the following pre-procedural reports: the two most recent discharge letters (including letters from external clinics), the invasive coronary angiography report, the echocardiography report, the CT scan report, and the HT protocol. We manually anonymized these reports. HT meeting protocols are standardized documents that contain decision-relevant patient characteristics, such as comorbidities, surgical risk scores, as well as the final treatment decision of the HT (Supplementary material, Figure S1). A detailed description of our institutional HT is provided in the Supplementary.

### Large Language Models

The study employed several state-of-the-art LLMs, namely GPT-3.5^11^, GPT-4^12^,GPT-4 Turbo, and GPT-4o by OpenAI, and PaLM 2 by Google^14^. In addition, we used the open-source models DeepSeek-R1^13^ by DeepSeek, Mistral-7B^15^, Llama 2 by Meta^16^ and BioGPT^17^. These LLMs had either demonstrated proficiency in similar tasks or had undergone pre-training on medical literature. Model details are provided in Supplementary Table S1. The model hyperparameters were set to the default values, except for the temperature, which was set to zero in accordance with previous studies in the medical domain.^18^ Temperature is a hyperparameter that controls the randomness of the LLM’s output. Lower values make the output more deterministic and focused, reducing variability and creativity. A detailed description of how we accessed the LLMs and handled input size constraints is given in the Supplementary material.

### Reference model

The reference model represented an algorithmic emulation of the 2021 ESC/EACTS Guidelines for the management of valvular heart disease.^10^ More specifically, the reference model assigned patients to either SAVR or TAVR according to a flowchart (Supplementary material, Figure S2) and relevant clinical variables (Table S4, Table S5).^10^ Model details are provided in the Supplementary.

### Experiments

Four experiments were conducted to investigate the effects of data pre-processing on LLM performance:

**RAW**: In the RAW experiment, anonymized medical reports (i.e., the two most recent discharge letters, the invasive coronary angiography report, the echocardiography report, and the CT scan report) were concatenated, and stored in a unified text file. This file was programmatically inserted into a prompt template. Each prompt included an introductory or continuation phrase and concluded with a request for a treatment decision (Supplementary material, Table S6).

**RAW+**: As it is unknown whether the LLMs we used had sufficient guideline knowledge, we compiled a resume of relevant CPG content from the ESC/EACTS guidelines.^10^ We added this resume to the prompt along with the unified text reports.

**SUM**: To study the effect of content compression, we replaced the original medical reports used in RAW with concise case summaries. These case summaries were created manually based on patient data extracted from the HT protocols.

**SUM+**: Case summaries were used as input and enriched with the CPG resume (Figure 1).

Prompt templates, the CPG resume and an exemplary case summary are shown in the Supplementary material (Table S6).

The LLMs’ responses were manually reviewed and categorized as either “TAVR”, “SAVR” or “indeterminate”. Indeterminate responses occur when the model output does not match the available answer choices or when the model determines that there is insufficient information to support a decision (Supplementary material, Table S7). To assess reliability and obtain robust estimates of performance metrics, the LLMs were presented with the same prompt input 10 times in succession for each experiment and patient (hereafter referred to as ‘runs’) to obtain a treatment decision. To prevent memory bias, a new chat session was initiated for each run.

### Performance metrics

We quantified agreement by means of Cohen’s kappa coefficients. For the sake of completeness, we also calculated accuracies as the proportion of treatment decisions that agreed with those made by the HT; however, we emphasize that due to class imbalance, this metric is only of limited significance and therefore only reported in the Supplementary. Decidability was quantified as the proportion of determinate treatment decisions. Bias was quantified using frequency bias indices (FBI), defined as the ratio of predicted to observed treatment decisions for TAVR.

Due to the limitations of individual metrics, we used two different metrics to quantify reliability: intraclass correlation coefficients (ICCs) and normalized Shannon entropy. A detailed description of the performance metrics, including strategies for handling indeterminate responses, is provided in the Supplementary material (Table S8).

### Statistical analysis

Patient characteristics for patients who received SAVR vs. TAVR were compared using Student’s t-test for normally distributed continuous variables and Mann-Whitney U test for non-normally distributed continuous variables. The Shapiro-Wilk test was used to assess normality. The chi-squared test was used for binary variables and Fisher’s exact test for sparse binary data.

Accuracy and Cohen’s kappa were computed with Python’s sklearn.metrics package (version 1.2.2). ICCs were calculated based on a one-way random effects, absolute agreement, single-rater model^19^ using Python’s pingouin package (version 0.5.3).

## Results

### Patient characteristics

A total of 80 patients with severe AS who were discussed at our institutional HT in 2022 were included. Of these patients, 24 (30 %) underwent SAVR while 56 (70 %) underwent TAVR. Patient characteristics are presented in Table 1.

**Table 1:** Patient characteristics. Values are mean ± standard deviation for continuous, normally distributed data, median and [interquartile range] for continuous, non-normally distributed data, and n (%) for binary data. EuroSCORE: European system for cardiac operative risk evaluation, SAVR: Surgical Aortic Valve Replacement, STS: Society of Thoracic Surgeons, TAVR: Transcatheter Aortic Valve Replacement

| Variable | With Data (%) | Overall | SAVR (n = 24) | TAVR (n = 56) | P-Value |
| --- | --- | --- | --- | --- | --- |
| Age (years) | 100.0 | 77.74 ± 7.5 | 70.71 ± 6.1 | 80.75 ± 5.8 | <0.001 |
| Female sex | 100.0 | 36 (45.0) | 8 (33.3) | 28 (50.0) | 0.26 |
| Height (cm) | 100.0 | 168.1 ± 11.0 | 172.5 ± 11.0 | 166.3 ± 10.6 | <0.05 |
| Body mass (kg) | 100.0 | 76.3 ± 17.0 | 79.0 ± 16.0 | 75.1 ± 17.4 | 0.35 |
| Body mass index (kg/m<sup>2</sup>) | 100 | 26.0 [23.0, 29.7] | 25.9 [23.2, 29.0] | 26.2 [23.0, 29.8] | 0.66 |
| Logistic EuroSCORE | 31.2 | 6.8 [4.5, 13.0] | 4.5 [2.2, 6.8] | 8.4 [5.0, 16.0] | 0.20 |
| EuroSCORE II | 98.8 | 2.6 [1.6, 4.5] | 1.8 [1.1, 3.1] | 2.9 [1.8, 4.9] | <0.05 |
| STS score | 76.2 | 2.8 [1.6, 4.5] | 1.4 [1.1, 3.0] | 3.3 [2.1, 4.5] | 0.12 |
| Left ventricular ejection fraction (%) | 100 | 60.0 [54.3, 61.3] | 60.0 [48.8, 62.0] | 60.0 [55.0, 60.0] | 0.28 |
| Aortic valve opening area (cm<sup>2</sup>) | 100 | 0.70 [0.60, 0.80] | 0.80 [0.68, 0.80] | 0.70 [0.60, 0.80] | 0.18 |
| Arterial hypertension | 100 | 59 (73.8) | 18 (75.0) | 41 (73.2) | 1.0 |
| Diabetes mellitus | 100 | 22 (27.5) | 6 (25.0) | 16 (28.6) | 0.96 |
| Hyperlipidemia | 100 | 51 (63.7) | 13 (54.2) | 38 (67.9) | 0.36 |
| Previous cardiac surgery | 100 | 1 (1.2) | 0 (0.0) | 1 (1.8) | 1.0 |
| Frailty | 100 | 7 (8.8) | 0 (0.0) | 7 (12.5) | 0.17 |
| Sequelae of chest radiation | 100 | 0 (0.0) | 0 (0.0) | 0 (0.0) | 1.0 |
| Porcelain aorta | 100 | 0 (0.0) | 0 (0.0) | 0 (0.0) | 1.0 |
| Expected patient-prosthesis mismatch | 100 | 1 (1.2) | 0 (0.0) | 1 (1.8) | 1.0 |
| Severe chest deformation or scoliosis | 100 | 7 (8.8) | 1 (4.2) | 6 (10.7) | 0.60 |
| Severe coronary artery disease requiring revascularization | 100 | 6 (7.5) | 5 (20.8) | 1 (1.8) | <0.05 |
| Left ventricular ejection fraction ≤ 40 % | 100 | 6 (7.5) | 3 (12.5) | 3 (5.4) | 0.52 |
| Active neoplasia | 100 | 7 (8.8) | 2 (8.3) | 5 (8.9) | 1.0 |
| Liver cirrhosis | 100 | 1 (1.2) | 0 (0.0) | 1 (1.8) | 1.0 |
| Chronic obstructive pulmonary disease (GOLD stage $\geq 3$) | 100 | 5 (6.2) | 1 (4.2) | 4 (7.1) | 1.0 |
| Pulmonary arterial hypertension | 100 | 8 (10.0) | 3 (12.5) | 5 (8.9) | 0.94 |
| Under immunosuppressive therapy | 100 | 10 (12.5) | 2 (8.3) | 8 (14.3) | 0.71 |

### Qualitative analysis

The LLMs’ output ranged from nonsensical treatment recommendations (e.g., heart transplant) and purely fabricated content, to correctly assessing the patient’s status, choosing the correct treatment option, and supporting the treatment decision with additional anatomical insights (Table 2).

**Table 2:** Representative responses from the Large Language Models (LLMs) The LLMs’ treatment responses included well-informed decisions but also hallucinations ranging from obvious misinformation to absurd treatment recommendations and logical errors. We largely adhered to the taxonomy for the description of hallucinations established by Huang et al.^27^ R: Response of the LLM with subscripts indicating responses to the same question (obtained during ten runs). Green: Correct or useful, red: incorrect or harmful. Abbreviations as in Table 1. * Exact patient ages are replaced by age ranges for this table to ensure anonymization. In the experiments the exact patient age in years was used.

| Model | Experiment | Patient characteristics | LLM response | HT treatment decision | Interpretation |
| --- | --- | --- | --- | --- | --- |
| PaLM2 | RAW | male, 55-59 years* old, EuroSCORE II: 0,55 %, no comorbidities except diffuse, mild coronary atherosclerosis and arterial hypertension, no relevant anatomical aspects to consider. | **R:** “Based on the provided information, the recommended treatment for the patient would be transcatheter aortic valve replacement (TAVR). The patient has severe aortic stenosis, which is causing symptoms and has been confirmed by echocardiography and cardiac catheterization. The patient is also at high risk for surgical aortic valve replacement (SAVR) due to their age, comorbidities, and the presence of diffuse coronary artery disease. TAVR is a less invasive procedure than SAVR and is associated with a lower risk of complications. Therefore, TAVR would be the preferred treatment option for this patient.” | SAVR | The LLM incorrectly recommends TAVR based on falsely extracted and/or interpreted patient characteristics (**factual inconsistency**).<br>According to the literature, it has not yet been conclusively determined whether TAVR is associated with fewer complications than SAVR<sup>37</sup> (**factual inconsistency/bias**). |
| BioGPT | RAW | male, 65-69 years* old, EuroSCORE- II: 7,2 %, post-cardiac arrest syndrome, frailty, long-term mechanical ventilation, liver cirrhosis, no relevant anatomical aspects to consider. | **R:** “Surgical replacement of the aortic valve. The patient was referred to the operating room for AVR. The operation was performed under general anesthesia and extracorporeal circulation ... The operation was performed with a median sternotomy. The ascending aorta was dissected free and opened...and the coronary arteries were visualized...” | TAVR | The LLM falsely states a surgery report from a procedure this patient never received and that was not mentioned in the prompt (**context inconsistency**). |
| BioGPT | RAW | female, 75-79 years * old, EuroSCORE- II: 2.4 %, STS score: 2.9%, COPD (GOLD Stage III), pulmonary hypertension, frailty, no relevant anatomical aspects to consider. | **R:** “Heart transplantation. - The patient's family decided against surgery. - The patient died due to heart failure.” | TAVR | The LLM did not follow the instruction to choose between SAVR and TAVR but opted for heart transplant (**instruction inconsistency**).<br>The LLM fabricated a clinical course that was not mentioned in the prompt (**context inconsistency**). |
| GPT35 | SUM | female, 70-74 years * old, EuroSCORE- II: 1.6 %, STS score: 1.1 %, no relevant comorbidities, no relevant anatomical aspects to consider. | **R1:** “Based on the given information, the recommended treatment for a person with severe aortic valve stenosis and no other significant comorbidities would be SAVR (Surgical Aortic Valve Replacement) if conservative therapy is not an option.”<br>**R2:** “Based on the provided information, the recommended treatment for a person with severe aortic valve stenosis, who does not | SAVR | The LLM gave opposite answers to the exact same question (**unreliability**).<br>The LLM incorrectly recommends TAVR based on the absence of comorbidities |
|  |  |  | have any contraindications or relevant comorbidities, would be TAVI (Transcatheter Aortic Valve Implantation) rather than SAVR (Surgical Aortic Valve Replacement)." |  | (factual inconsistency). |
| PaLM2 | RAW+ | male, 55-59 years * old, EuroSCORE- II: 0.55 %, STS score: 0.7 %, no comorbidities except arterial hypertension, no relevant anatomical aspects to consider. | R: "Based on the guidelines, the recommended treatment would be transcatheter aortic valve replacement (TAVR). The patient is 55-59 years old and at high surgical risk (STS-PROM score >8 % or EuroSCORE II > 8 %) due to severe frailty and previous cardiac surgery. TAVR is the preferred treatment for patients who are inoperable or at high surgical risk." | SAVR | The LLM falsely claims that the patient is at high surgical risk due to frailty and previous cardiac surgery – which are both entirely fabricated (factual inconsistency). |
| GPT35 | SUM+ | female, 80-84 years * old, logistic EuroSCORE: 8.44 %, EuroSCORE II: 1.82 %, STS score: 4.33 %, stage 3A chronic kidney disease, no relevant anatomical aspects to consider. | R: "Based on the guidelines provided, the recommended treatment for the patient would be SAVR (surgical aortic valve replacement) if conservative therapy is not an option. This is because the patient is 80-84 years old and falls into the older age category, and their logistic EuroSCORE is 8.44 %, which is above the threshold for TAVR (transcatheter aortic valve replacement) recommendation." | TAVR | The LLM incorrectly recites the association between the logistic EuroSCORE and surgical risk prohibiting SAVR (context inconsistency).<br>The LLM correctly assigns the patient to the older age category but then incorrectly recommends SAVR as the optimal therapy (logical inconsistency). |
| GPT4 | RAW | female, 65-69 years * old, EuroSCORE- II: 2.5 %, STS score: 1.4 %, no relevant comorbidities, ascending aortic aneurysm (48 mm) mentioned in the CT scan report. | R: "Based on the guidelines, the recommended treatment would be Surgical Aortic Valve Replacement (SAVR) as the patient has a severe aortic stenosis and an ascending aortic aneurysm. Transcatheter Aortic Valve Implantation (TAVI) is typically recommended for patients who are at high or prohibitive surgical risk, which does not seem to be the case here." | SAVR | Correct treatment decision plus additional extraction of decision-relevant clinical data. |
| LLaMA2 | SUM+ | male, 65-69 years * old, EuroSCORE- II: 1.29 %, STS score: 3.04 %, COPD, no relevant anatomical aspects to consider. | R1: "Based on the guidelines provided, the recommended treatment for this patient would be TAVI. The patient's age (65-69 years old) and low surgical risk (EuroSCORE II 1.29 %, STS score 3.037 %) make them a suitable candidate for TAVI. Additionally, the patient does not have any contraindications for TAVI, such as severe frailty, previous cardiac | SAVR | The LLM correctly identifies low surgical risk but then incorrectly rejects SAVR as therapeutic option (logical inconsistency).<br>The LLM incorrectly recites factors favoring TAVR from |
|  |  |  | surgery, or significant thorax deformity.”<br>**R<sub>2</sub>:** “Based on the information provided, the patient's EuroSCORE II is 1.29 % and STS score is 3.037 %, which indicates that they are at low surgical risk. The patient is also younger than 75 years old. Therefore, according to the guidelines, SAVR is the recommended treatment option for this patient.” |  | the prompt (**context inconsistency**).<br>Although the LLM correctly decided for SAVR this time, it gave the opposite response to the exact same question (**unreliability**). |
| DeepSeek-R1 | RAW+ | male, 65-69 years* old, EuroSCORE II: 0.92%, STS score: 0.73%, end-stage renal disease requiring hemodialysis, no relevant anatomical aspects to consider. | **R:** “... Based on the 2021 ESC/EACTS guidelines and the patient's clinical profile, the recommended treatment would be TAVR.<br>Reasoning:<br>• Patient age (65-69 years) is below 75 but has significant comorbidities, including terminal renal insufficiency, indicating high surgical risk.<br>• Suitable tricuspid aortic valve anatomy for TAVR.<br>• High surgical risk factors favor TAVR over SAVR.” | SAVR | The LLM overestimates the effect of end-stage renal disease on surgical risk, which was still relatively low overall, as evidenced by low surgical risk scores (**factual inconsistency**). |

Qualitative analysis revealed that smaller models (e.g., BioGPT) tended to provide conflicting treatment recommendations for the same patient. In contrast, the frontier models (e.g., GPT-4, PaLM 2) consistently provided the same treatment recommendation when presented with the same patient data repeatedly over 10 runs.

In each experiment, all LLMs produced hallucinations of varying severity and frequency. These included instructional, contextual and factual inconsistencies (Table 2).

### Quantitative analysis

Figure 2 and Table S9 (Supplementary material) present the performance metrics. In the RAW experiment, LLMs’ treatment decisions were in poor agreement with the HT. In this experiment, DeepSeek-R1 showed the highest agreement with the HT with a Cohen’s kappa coefficient of 0.22. Some LLMs gave indeterminate treatment recommendations in up to 54 % of cases (e.g., GPT-3.5) and showed low reliability as evidenced by low ICCs and high entropy values (e.g., Mistral, Llama-2 and DeepSeek-R1). FBIs were substantially higher than 1.0 for all LLMs except BioGPT, indicating a bias towards TAVR. The reference model outperformed the LLMs in the RAW experiment regarding the metrics we assessed.

**Figure 2:**
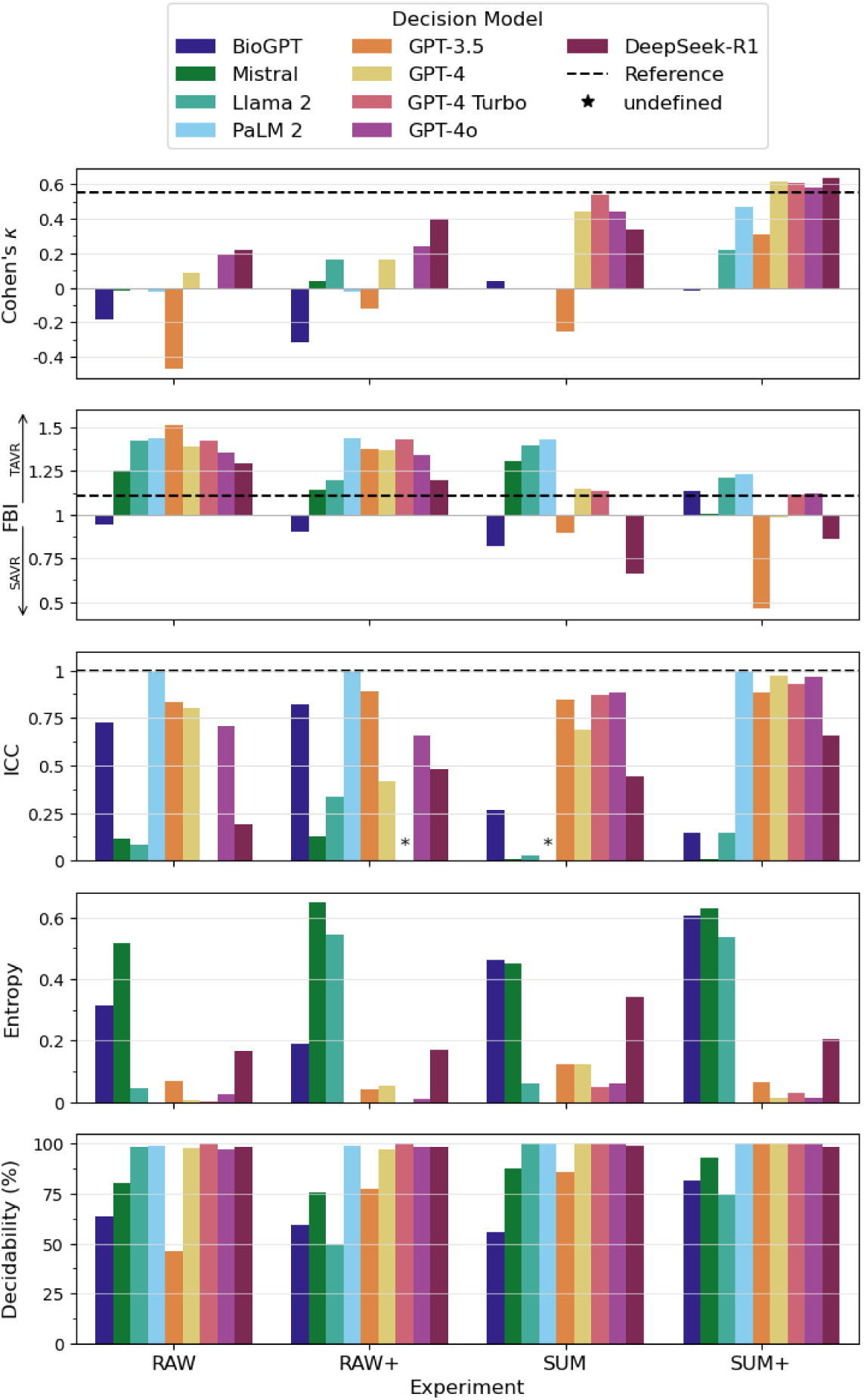
Performance metrics. Performance metrics of the Large Language Models (LLMs) are shown for the four experiments we conducted. The dashed line represents the reference model. Cohen’s kappa coefficients ≤ 0 indicate no agreement, 0.01-0.20 slight, 0.21-0.40 fair, 0.41-0.60 moderate, 0.61-0.80 substantial, and 0.81-1.0 almost perfect agreement^37^ with the Heart Team’s treatment decisions. Frequency bias indices (FBIs) > 1 indicate bias towards TAVR and < 1 bias towards SAVR. Intraclass correlation coefficients (ICCs) < 0.5 indicate poor, 0.50-0.75 moderate, 0.75-0.90 good, > 0.90 excellent test-retest reliability.^19^ Instances where ICCs were undefined are marked by asterisks. Entropy values close to zero indicate low and entropy values close to one indicate high output variation, respectively. Decidability was defined as the proportion of non-indeterminate treatment decisions. The exact numerical values for the performance metrics are displayed in the Supplementary material (Table S9). Abbreviations as in Figure 1.

In the RAW+ experiments, DeepSeek-R1 again showed the highest agreement with the HT with a Cohen’s kappa coefficient of 0.40 indicating fair agreement. The performance metrics of the other LLMs did not change substantially in the RAW+ experiment. However, the performance metrics of most LLMs substantially improved in the SUM experiment and peaked in the SUM+ experiment, where some LLMs (e.g., GPT-4 models and DeepSeek-R1) drew level with the reference model.

A general trend towards more concordant treatment decisions, fewer indeterminate responses, increased reliability, and less bias towards TAVR was observed with increasing data pre-processing and information enrichment efforts from RAW to SUM+ (Figure 2, Figure 3).

**Figure 3:**
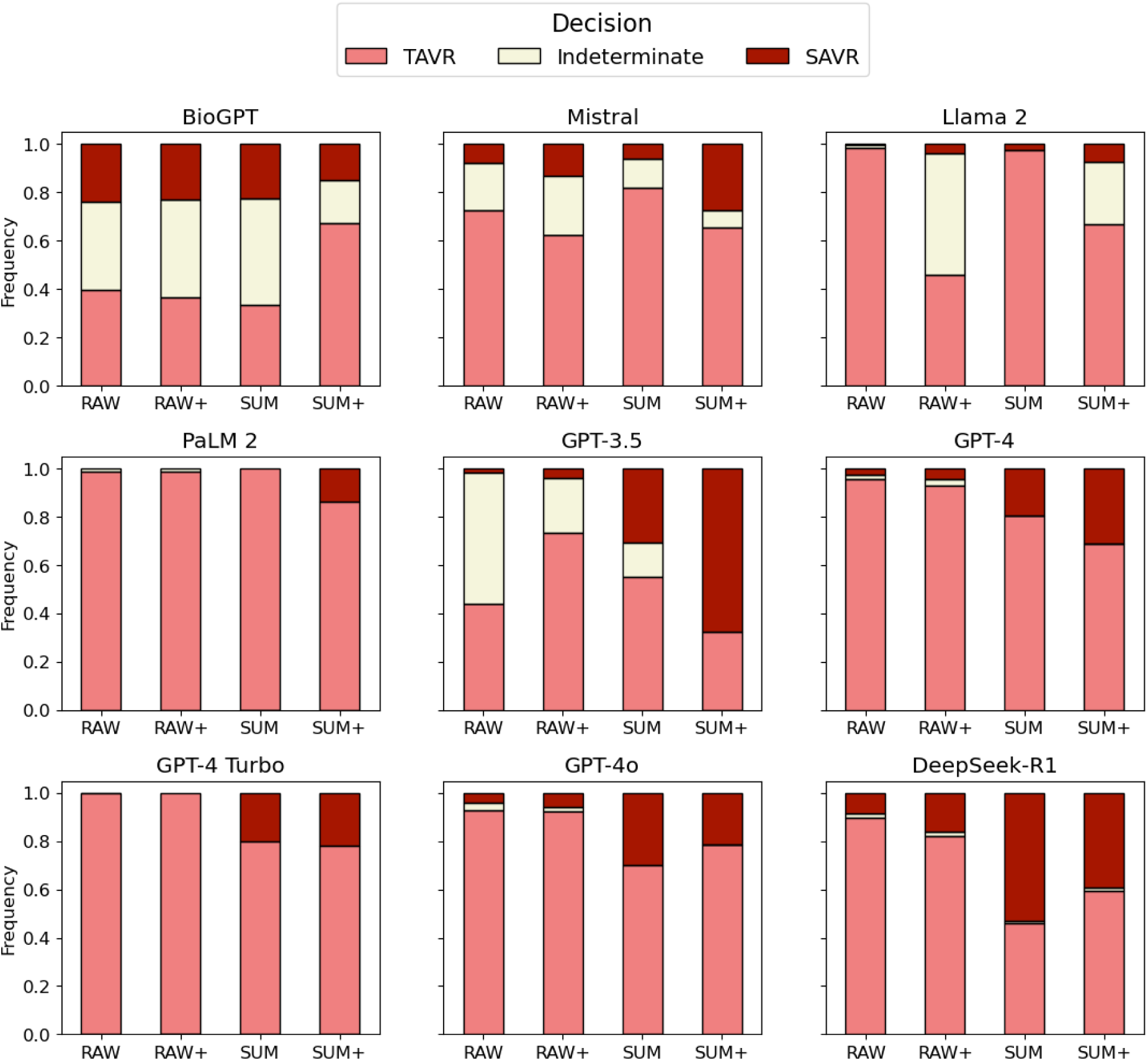
Frequencies of treatment decisions. Frequency distributions of the Large Language Models’ treatment decisions are shown. A general trend towards increasing decidability and an increasing proportion of treatment decisions favoring SAVR could be observed between the RAW and the SUM+ experiment. Abbreviations as in Figure 2.

## Discussion

To our knowledge, this is the first study to evaluate the impact of input data representation, including real-world medical data, on the ability of LLMs to make guideline-concordant treatment decisions.

### Current LLMs make incorrect decisions based on original clinical data

Our analysis reveals that LLMs struggled to process original medical reports effectively, often outputting ‘TAVR’ or providing indeterminate responses. The LLMs showed low agreement with the HT, exhibited undecidability and unreliability, and displayed a strong bias towards TAVR. The considerably high accuracies (Table S9) observed with some LLMs in the RAW experiment can be largely attributed to the class imbalance within our patient cohort, where 70 % of patients received TAVR.

### LLMs require extensive data pre-processing to make sound therapeutic decisions

Performance improved substantially when physician-made case summaries were used as input, and when guideline knowledge was added to the prompts. The GPT-4 models and DeepSeek-R1 stood out as the most capable LLMs in our experiments. When given case summaries and a CPG resume, these two models showed substantial agreement with HT and drew level with the reference model in terms of interrater agreement, decidability, and bias.

### Data representation affects LLM performance

GPT-4o, a distilled and streamlined version of GPT-4, and DeepSeek-R1, a model with enhanced reasoning abilities, showed more promising results than previous-generation LLMs when confronted with real-world medical data (RAW/RAW+); however, their performance remains insufficient for clinical application. The fact that even state-of-the-art LLMs show significant stochastic variations in decision-making – and thus unreliability – further supports this finding.

An explanation for the underperformance of LLMs in the RAW experiment is not immediately apparent due to their opaque nature and the lack of established tools that allow the direct examination of input-output correlations. However, the underperformance cannot be attributed to a lack of or incorrectly applied guideline knowledge since the performance in RAW+ was in general similar to RAW and since LLMs can presumably apply clinical knowledge to clinical cases as shown in their ability to pass medical board exams.^1,20^ This, along with the significant performance gains observed when providing case summaries instead of original medical reports, suggests that input data representation is the most critical factor in LLM performance.

This finding is consistent with the fact that virtually all studies in which LLMs have been shown to make sound treatment decisions, used pre-processed clinical data as input.^4–8^ Of note is the study by Salihu et al.^8^ In this study, data from patients with severe AS were provided to GPT-4 to obtain a treatment decision for either TAVR, SAVR or conservative management. Patient data were provided in the form of a standardized multiple-choice questionnaire with 14 key clinical variables as input, similar to our SUM experiments. GPT-4 treatment decisions were in substantial agreement with HT treatment decisions, a finding that we were able to reproduce in our experiments.

Similarly, in studies on tasks beyond therapeutic decision-making, such as answering board exam questions^1^ and diagnosing complex clinical cases^2,21^, LLMs performed particularly well when the input data were concise and information-dense.

Basic research has indicated that LLMs struggle with lengthy texts^22^ spanning over multiple prompts, potentially leading to memory loss^23^ and texts with a low signal-to-noise ratio.^24^ A study by Levy et al^25^ demonstrated that LLM reasoning performance declined notably with increasing input length. Specifically, the authors observed a 26 % drop in LLM performance when the input length was artificially increased from 250 to 3,000 tokens – i.e. a range of input lengths comparable to our study (see Supplementary material Table S3).

Recently, Hager et al.^26^ investigated the ability of LLMs to correctly diagnose patients presenting to the emergency department with abdominal pain. In this study, it was shown that deliberately withholding relevant clinical information from the LLMs paradoxically improved their diagnostic accuracy. Overall, this implies that LLMs are sensitive to both the signal-to-noise ratio and the sheer quantity of information provided.

### LLMs are not yet ready for clinical decision-making

The results obtained with pre-processed patient data in our study and in previous studies demonstrate the potential of LLMs in medicine. However, the use of curated and pre-processed data does not reflect the clinical situation: To this day, the communication of clinical data within hospitals is largely based on unstructured free text.

Healthcare professionals have high expectations of AI to reduce their workload. This is not the case when physicians must manually extract and prepare key patient data for LLMs, as data extraction, not the actual decision-making task, is usually the most labor-intensive step.

Once key patient data has been extracted and prepared as input, simpler machine learning models (e.g., tree-based models) could be used alternatively to provide decision support. In our study, as well as in Salihu et al.’s^8^ study, simple reference models performed comparably to GPT-4, suggesting that non-LLM models could outperform LLMs if trained appropriately. In addition, non-generative models do not exhibit undesirable behaviors such as hallucinations and unreliability^27,28^, and provide explainability and established measures of uncertainty quantification, two hallmarks of reasonable AI^29^ that are currently not adequately implemented for LLMs^30,31^.

Another hallmark of reasonable AI is to address algorithmic bias. It is conceivable that the bias we observed in virtually all LLMs in our study could be due to LLMs being exposed to an abundance of TAVR-related internet literature during training compared to SAVR, subsequently influencing their treatment decisions.

A reasonable approach could be to use LLMs to extract clinical data^32^ and generate input for downstream non-LLMs, which then perform the decision-making. While this strategy should ideally exploit the strengths of LLMs and well-established machine learning classifiers, its effectiveness remains to be proven in future studies.

### Limitations

Our study is subject to certain limitations. These include a small patient cohort from a single center and the retrospective nature of our investigation. Nevertheless, the size of our study cohort (n=80), was comparable to previous key publications^2,33^ studying the performance of LLMs in medicine, and we assume that our patient cohort was sufficiently large given the clear trends we observed.

The HT decisions against which we compared the LLMs’ treatment decisions may themselves be non-objective and deviate from the CPGs. We manually reviewed the HT treatment decisions and found no substantial deviations from the CPGs. Since treatment decisions are ultimately made by a team of physicians, i.e., human individuals, the ground truth in experiments such as ours is inherently susceptible to some degree of subjectivity.

Given the limited cohort size and the considerable length of the medical reports, few-shot prompting or fine-tuning was not a viable option. We did not employ more sophisticated prompting techniques, such as Chain-of-Thought^35^, and confined hyperparameter tuning to the temperature parameter.

## Conclusions

Our experiments have been among the most challenging tasks LLMs have been asked to perform in the medical sciences. Overall, we conclude that LLMs are currently not suitable as decision makers for the treatment of patients with severe AS, as our results suggest that LLMs require elaborate pre-processing of patient data to make guideline-concordant treatment decisions. Thus, we do not share the medical community’s concern that staff will be replaced by artificial intelligence^36^ in clinical decision-making in the near future.

Our findings suggest that LLMs should be used cautiously, particularly by medical laypersons seeking medical advice, such as second opinions. Users without extensive domain knowledge may receive treatment recommendations at a level similar to our RAW experiments. This is because medical laypersons may not be able to support prompts with guideline knowledge or create case summaries of sufficient quality but will only be able to use original medical reports. The study by Hager et al.^26^ suggests that LLMs perform poorly when collecting additional patient data sequentially, as physicians would during a patient-physician dialogue. This suggests that the alternative to our approach – not providing all clinical data to the LLM at once, but having medical laypersons provide essential information incrementally during a chat session - is also likely to lead to suboptimal therapeutic recommendations.

Finally, medical laypersons may not be able to recognize hallucinations as effectively as medical professionals. This, combined with the eloquent and persuasive linguistic style of most LLMs, has the potential to mislead users by creating an illusion of greater certainty than warranted, aggravating the hazardous effects of incorrect treatment recommendations.

## Data Availability

All data produced in the present study are available upon reasonable request to the authors

## Acknowledgements

Dr. Roeschl and Dr. Hashemi are participants in the BIH Charité Digital Clinician Scientist Program funded by the Charité – Universitätsmedizin Berlin, and the Berlin Institute of Health at Charité (BIH).

We thank Michael Gudo (MORPHISTO GmbH) for providing access to GPT-4 and Hadi El Ali (B.Sc.), University of Bayreuth, for contributing to the illustration of Figure 1.

## Conflicts of Interests

Djawid Hashemi reports financial engagements beyond the scope of the presented work. These activities include consultation services and speaking engagements for companies including AstraZeneca, Bayer Vital, Boehringer Ingelheim, Coliquio, and Novartis.

Tobias D. Trippel reports on the potential conflict of interest by holding shares of Microsoft, Amazon and Palantir Technologies.

Axel Unbehaun serves as physician proctor to Boston Scientific, Edwards Lifesciences, and Medtronic.

Jörg Kempfert reports personal fees from Edwards, personal fees from LSI, outside the submitted work.

Benjamin O’Brien declares Research funding from the British Heart Foundation and the National Institute for Health Science Research and relevant financial activities outside the submitted work with following commercial entities: Teleflex, Abiomed in relation to consultancy fees.

Felix Balzer reports funding from Medtronic and grants from the German Federal Ministry of Education and Research, grants from the German Federal Ministry of Health, grants from the Berlin Institute of Health, personal fees from Elsevier Publishing, grants from Hans Böckler Foundation, other from Robert Koch Institute, grants from Einstein Foundation, and grants from Berlin University Alliance outside the submitted work.

Volkmar Falk declares relevant financial activities outside the submitted work with following commercial entities: Medtronic GmbH; Biotronik SE & Co.; Abbott GmbH & Co. KG; Boston Scientific; Edwards Lifesciences; Berlin Heart; Novartis Pharma GmbH; JOTEC GmbH; Zurich Heart. In relation to: Educational Grants (including travel support); Fees for lectures and speeches; Fees for professional consultation; Research and study funds.

Alexander Meyer declares the receipt of consulting and lecturing fees from Medtronic, lecturing fees from Bayer, and consulting fees from Pfizer. Alexander Meyer is founder and managing director of x-cardiac GmbH.

The remaining authors have no conflicts of interest to disclose.

## Data sharing statement

The (anonymized) data underlying this article will be shared on reasonable request to the corresponding author.

## Author Contributions

Conception and design of the study and literature review: TR, MH, DH, AM. Data collection: DH, FR. Analysis and interpretation of the data: MH, TR, AM, NH, MG. Drafting of the manuscript: TR, MH, DH, AM, NH. All authors: revising and editing the manuscript.

## Supplementary methods

### Access to LLMs

We accessed the GPT models and PaLM 2 using the application programming interfaces (API) of OpenAI and Google, respectively. Mistral, DeepSeek-R1 and Llama 2 were accessed through the REST API of perplexity. BioGPT was downloaded from Huggingface and run locally.^1^

For all LLMs except BioGPT and Mistral, we were able to clearly demonstrate that the LLMs were familiar with the 2021 ESC/EACTS Guidelines for the management of valvular heart disease^2^ and could correctly recite key contents.

### Language

All medical reports were originally available in German and were fed into the LLMs as such except for BioGPT. Since BioGPT has not been sufficiently trained on German text data, we translated the original medical reports to English using Python’s deep-translator (version 1.11, module: GoogleTranslator) for these experiments.

### Handling of input size constraints

When the model-specific text input sizes were exceeded, we broke up the text into chunks and modified the prompt to inform the model about contiguous patient data. We established context by adding the preceding output. The context sizes are given in Table S1 and range from 1,024 to 128,000 tokens. The median token counts and average number of prompts are shown in Table S2 and Table S3.

### Institutional Heart Team

Our institutional Heart Team (HT) is comprised of interventional cardiologists, cardiac surgeons, imaging specialists and cardiac anesthesiologists. It is mandatory that at least one representative of the aforementioned specialties participates in an HT meeting. HT meetings are held every two weeks to make treatment decisions for patients with coronary artery disease, valve disorders and structural heart disease.

The HT follows a structured decision-making process outlined in the HT protocol (Figure S1). In this process, decision-relevant patient data (e.g., patient characteristics, comorbidities, anatomical aspects) previously extracted from discharge letters and diagnostic imaging reports is presented along with imaging scans to the HT members. Surgical risk scores are calculated and documented in the HT protocol along with decision-relevant patient data. Patients are assigned to the respective treatment modality according to a guideline-guided approach as depicted by the structure of the HT protocol.

For patients in our study cohort, the ESC/EACTS Guidelines for the management of valvular heart disease^2^, published online in August 2021, were used as the basis for decision-making. The members of the Heart Team are collectively committed to follow these guidelines. If a patient requests a different therapy from that recommended by the Heart Team, this is documented in the HT protocol. In our study, the therapy decisions made by the LLMs were compared to the primary therapy recommendations by the HT, regardless of the patient’s preferences. This was the case for one out of 80 patients in our study cohort.

### Reference model details

The reference model represented an algorithmic emulation of the ESC/EACTS Guidelines for the management of valvular heart disease.^2^ The reference model consisted of a decision tree combined with a weighted sum model (WSM). The decision tree assigned patients to either SAVR or TAVR according to the flowchart outlined in Figure S2. For patients who could not be unambiguously assigned to one or the other treatment modality according to the flowchart, we applied the WSM to arrive at a treatment decision. The WSM linearly combined decision-relevant clinical variables *v* and corresponding weights *w* to arrive at a WSM score *S*:

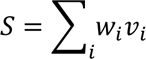

*S* ≤ 0 indicated a decision for SAVR and a *S* > 0 indicated a decision for TAVR. The input data for the reference model was extracted from the HT protocols. The WSM included variables listed in the ESC/EACTS guidelines^2^ shown in Table S4. The variable weights were determined via a consensus-seeking process among cardiologists and cardiac surgeons (Table S5). Performance metrics were calculated accordingly for the reference model, except that the ICC was set to 1 and the entropy set to 0 due to the purely deterministic nature of the reference model.

## Supplementary Figures

**Figure S1:**
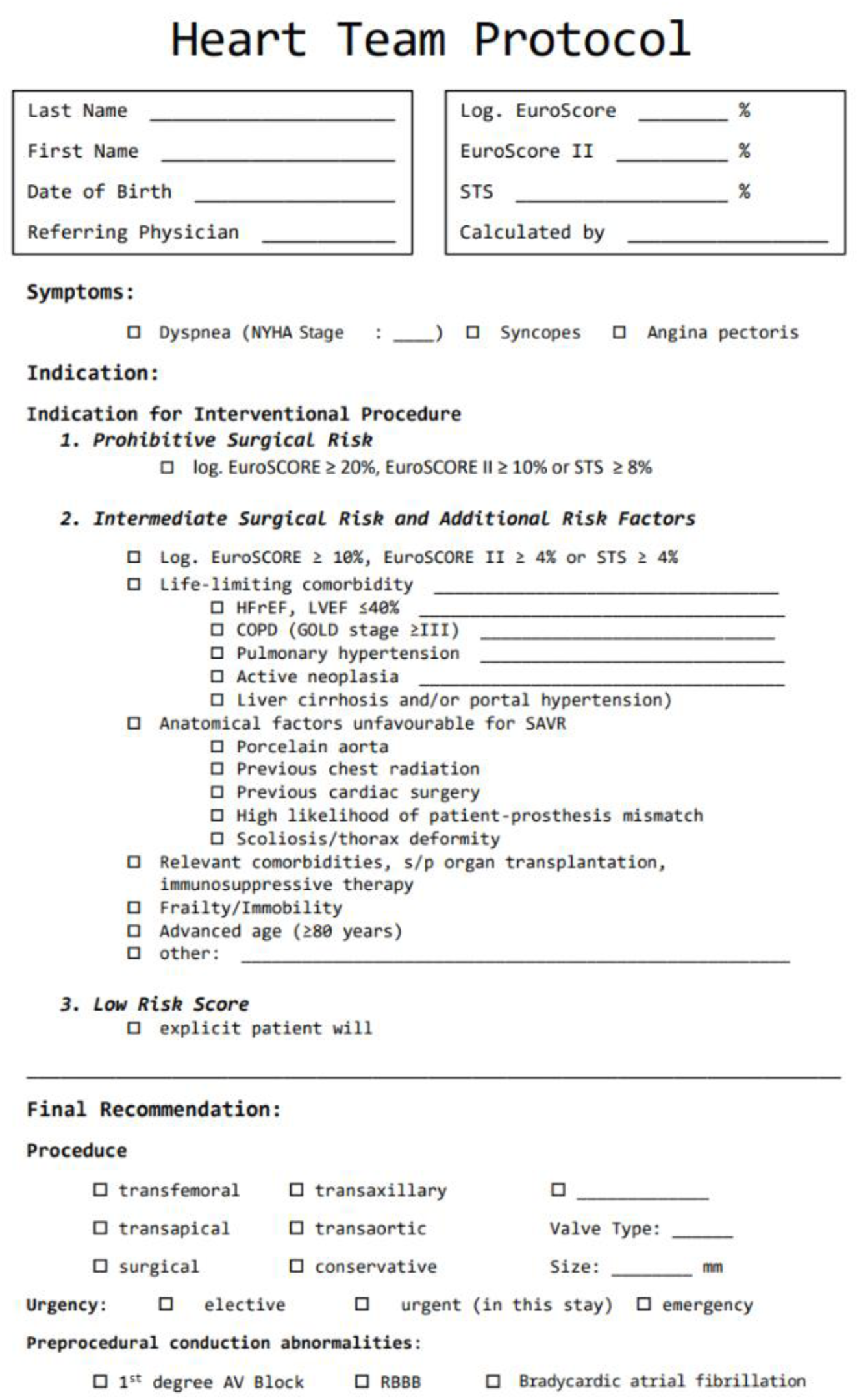
Heart Team Protocol. Our institutional Heart Team (HT) protocols included the HT’s treatment decision in addition to decision-relevant patient characteristics. These patient characteristics were used to create case summaries for the SUM and SUM+ experiments and were used as input for the reference model. RBBB: Right bundle branch block. Abbreviations as in Tables S1-2.

**Figure S2:**
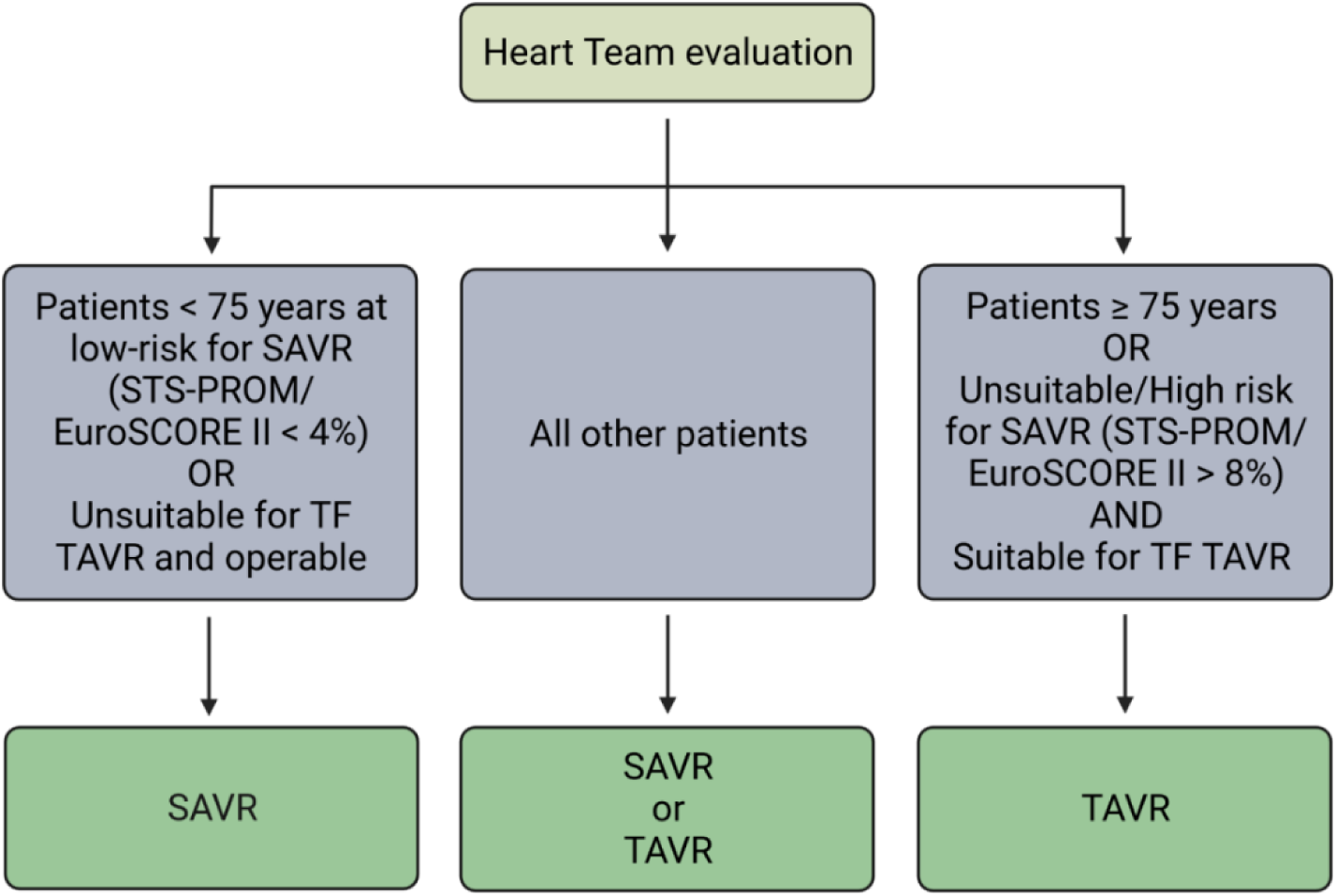
Flowchart for the management of patients with severe AS. Management of patients with severe aortic stenosis (AS) according to the 2021 ESC/EACTS Guidelines for the management of valvular heart disease.^2^ TF: Transfemoral. Other abbreviations as in Tables S1-2.

## Supplementary Tables

**Table S1:** Model cards. Model version refers to the identifiers used for each Application Programming Interface (API). Model size is given as the estimated number of trainable parameters. The context size refers to the maximum number of tokens per prompt. Note that detailed model sizes, architectures, training sets and methods are not disclosed by OpenAI and Google and are listed without warranty for correctness. Details on the training data of pre-trained and fine-tuned Llama 2 models are also not publicly available.

| Model Name | Version | Model Size<br>(parameters) | Pre-Training | Context Size<br>(tokens) |
| --- | --- | --- | --- | --- |
| BioGPT | microsoft/BioGPT-Large-PubMedQA | $359 \times 10^6$ | large scale biomedical literature | 1,024 |
| ChatGPT-3.5 | gpt-3.5-turbo-0613 | $175 \times 10^9$ | $500 \times 10^9$ tokens including books, Wikipedia, academic papers, and WordPress-based websites until September 2021. Fine-tuned via reinforcement learning with human feedback (RLHF). | 4,096 |
| ChatGPT-4 | gpt-4-0613 | $\sim 1\text{--}1.8 \times 10^{12}$ based on 16 models with 110 billion parameters each, connected by a Mixture of Experts (MoE) | $13 \times 10^{13}$ tokens including data from CommonCrawl, RefinedWeb, and social media until September 2021, with some select information from beyond that date. The model was fine-tuned with data from ScaleAI and internal sources, and RLHF. | 8,192 |
| ChatGPT-4 Turbo | gpt-4-1106-preview | $\sim 1.8 \times 10^{12}$ based on 16 models with $\sim 111$ billion parameters each, connected by a Mixture of Experts (MoE) | $13 \times 10^{13}$ tokens including data from CommonCrawl, RefinedWeb, and social media until September 2021, with some select information from beyond that date. The model was fine-tuned with data from ScaleAI and internal sources, and RLHF. | 128,000 |
| ChatGPT-4o | gpt-4o | $\sim 200 \times 10^9$ | Multimodal dataset including text, audio, and visual data with text data from publicly available internet data and licensed third-party datasets, and RLHF. | 128,000 |
| Llama 2 | Llama 2-70b-chat | $73 \times 10^9$ | $2 \times 10^{12}$ tokens of data from publicly available sources between January and July 2023 | 4,096 |
|  |  |  | SFT on publicly available instruction datasets and RLHF on over one million human-annotated examples. |  |
| Mistral | mistral-7b-instruct | $7 \times 10^9$ | Pre-trainset unknown, fine-tuned using a variety of publicly available conversation datasets. | 4,096 |
| PaLM 2 | text-bison-32k | $540 \times 10^9$ | Web documents, books, source code, mathematics, and conversational data | 32,000 |
| DeepSeek-R1 | sonar-reasoning | $671 \times 10^9$ MoE system with $21 \times 10^9$ parameters per expert | reasoning-specific tasks generated by DeepSeek-V3-Bases, synthetic data, RL with rewards derived from automated checks | 128,000 |

**Table S2:** Number of prompts per model and experiment. For each model and experiment the average number of prompts needed to present a complete patient case are shown. The number of prompts is contingent upon the context window size (Table S1) and the model-specific tokenizers.

| Model | RAW | RAW+ | SUM | SUM+ |
| --- | --- | --- | --- | --- |
| BioGPT | 6.4 | 6.6 | 1.0 | 1.0 |
| GPT-3.5 | 2.14 | 2.24 | 1.0 | 1.0 |
| GPT-4 | 1.18 | 1.27 | 1.0 | 1.0 |
| GPT-4 Turbo | 1.0 | 1.0 | 1.0 | 1.0 |
| GPT-4o | 1.0 | 1.0 | 1.0 | 1.0 |
| Llama 2 | 1.85 | 2.0 | 1.0 | 1.0 |
| Mistral | 1.7 | 1.8 | 1.0 | 1.0 |
| PaLM 2 | 1.0 | 1.0 | 1.0 | 1.0 |
| DeepSeek-R1 | 1.0 | 1.0 | 1.0 | 1.0 |

**Table S3:**
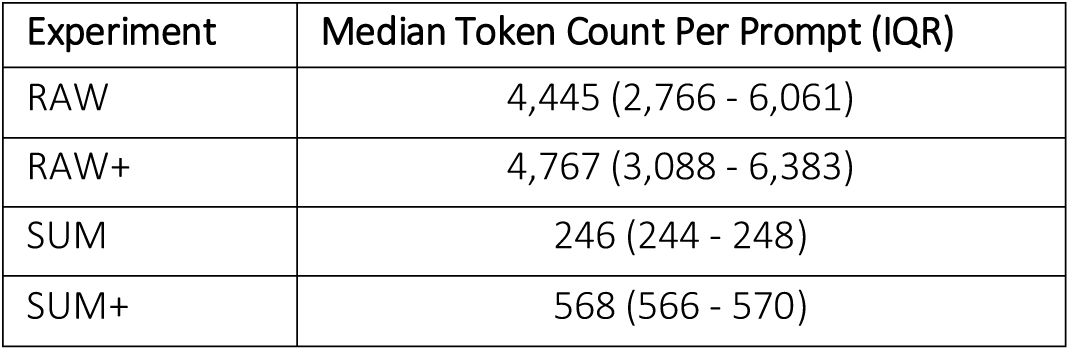
Token counts. For each experiment, median token counts per prompt are reported with interquartile ranges (IQR). Abbreviations as in Tables S1-2.

**Table S4:** Decision-relevant clinical variables according to the 2021 ESC/EACTS Guidelines for the management of valvular heart disease. Decision-relevant clinical variables favoring either SAVR or TAVR according to the 2021 ESC/EACTS Guidelines for the management of valvular heart disease.^2^ AVA: aortic valve area, BSA: body surface area, CAD: coronary artery disease, LVOT: Left ventricular outflow tract. Other abbreviations as in Figure S1.

|  | Favors<br>TAVR | Favors<br>SAVR |
| --- | --- | --- |
| **Clinical characteristics** |  |  |
| Lower surgical risk | – | + |
| Higher surgical risk | + | – |
| Younger age | – | + |
| Older age | + | – |
| Previous cardiac surgery (particularly intact coronary artery bypass grafts at risk of injury during repeat sternotomy) | + | – |
| Severe frailty | + | – |
| Active or suspected endocarditis | – | + |
| **Anatomical and procedural factors** |  |  |
| TAVR feasible via transfemoral approach | + | – |
| Transfemoral access challenging or impossible and SAVR feasible | – | + |
| Transfemoral access challenging or impossible and SAVR inadvisable | + | – |
| Sequelae of chest radiation | + | – |
| Porcelain aorta | + | – |
| High likelihood of severe patient-prosthesis mismatch ($AVA < 0.65 \text{ cm}^2/\text{m}^2$) | + | – |
| Severe chest deformation or scoliosis | + | – |
| Aortic annular dimensions unsuitable for available TAVR devices | – | + |
| Bicuspid aortic valve | – | + |
| Valve morphology unfavourable for TAVR (e.g., high risk of coronary obstruction due to low coronary ostia or heavy leaflet/LVOT calcification) | – | + |
| Thrombus in aorta or LV | – | + |
| **Concomitant cardiac conditions requiring intervention** |  |  |
| Significant multi-vessel CAD requiring surgical revascularization | – | + |
| Severe primary mitral valve disease | – | + |
| Severe tricuspid valve disease | – | + |
| Significant dilatation/aneurysm of the aortic root and/or ascending aorta | – | + |
| Septal hypertrophy requiring myectomy | – | + |

**Table S5:** Variables of the weighted sum model. Variables and variable weights of the weighted sum model (WSM) are shown. The WSM was applied to arrive at a treatment decision for patients who could not be unambiguously assigned to either surgical- or transcatheter aortic valve replacement according to the flowchart shown in Figure S2. The variable weights were determined via a consensus-seeking process among cardiologists and cardiac surgeons. BMI: body mass index. Other abbreviations as in Table S4.

| Variables from our institutional HT protocol | Weights |
| --- | --- |
| Severe CAD requiring surgical revascularization | -1 |
| Left ventricular ejection fraction < 40 % | 1 |
| COPD | 1 |
| Pulmonary arterial hypertension | 1 |
| Active neoplasia | 1 |
| Liver cirrhosis | 1 |
| Porcelain aorta | 5 |
| Sequelae of chest radiation | 1 |
| Previous cardiac surgery | 1 |
| Expected patient-prosthesis mismatch | 1 |
| Severe chest deformation or scoliosis | 1 |
| Under immunosuppressive therapy | 1 |
| Frailty | 5 |
| Cachexia (BMI < 18.5 kg/m<sup>2</sup>) or morbid obesity (BMI ≥ 40 kg/m<sup>2</sup>) | 1 |

**Table S6:**
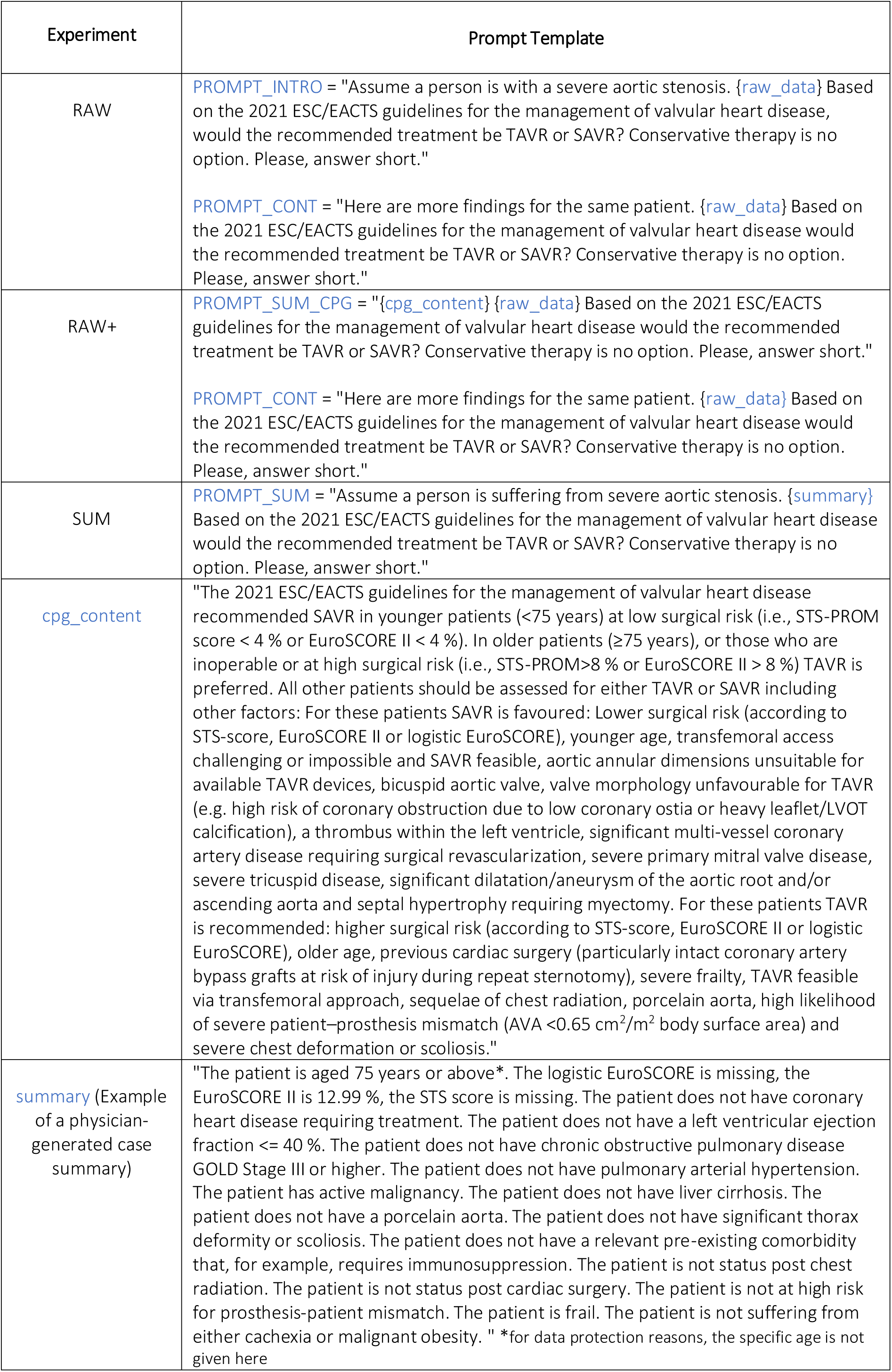
Prompt templates. Prompt templates used to communicate with the LLMs. The prompt templates were filled with the original medical reports (RAW) or case summaries (SUM) and/or a resume of the ESC/EACTS guidelines^2^ positionally indicated by the variables raw_data, summary and cpg_content, respectively. Abbreviations as in Figure S4.

| Experiment | Prompt Template |
| --- | --- |
| RAW | **PROMPT_INTRO** = "Assume a person is with a severe aortic stenosis. {raw_data} Based on the 2021 ESC/EACTS guidelines for the management of valvular heart disease, would the recommended treatment be TAVR or SAVR? Conservative therapy is no option. Please, answer short."<br>**PROMPT_CONT** = "Here are more findings for the same patient. {raw_data} Based on the 2021 ESC/EACTS guidelines for the management of valvular heart disease would the recommended treatment be TAVR or SAVR? Conservative therapy is no option. Please, answer short." |
| RAW+ | **PROMPT_SUM_CPG** = "{cpg_content} {raw_data} Based on the 2021 ESC/EACTS guidelines for the management of valvular heart disease would the recommended treatment be TAVR or SAVR? Conservative therapy is no option. Please, answer short."<br>**PROMPT_CONT** = "Here are more findings for the same patient. {raw_data} Based on the 2021 ESC/EACTS guidelines for the management of valvular heart disease would the recommended treatment be TAVR or SAVR? Conservative therapy is no option. Please, answer short." |
| SUM | **PROMPT_SUM** = "Assume a person is suffering from severe aortic stenosis. {summary} Based on the 2021 ESC/EACTS guidelines for the management of valvular heart disease would the recommended treatment be TAVR or SAVR? Conservative therapy is no option. Please, answer short." |
| cpg_content | "The 2021 ESC/EACTS guidelines for the management of valvular heart disease recommended SAVR in younger patients (<75 years) at low surgical risk (i.e., STS-PROM score < 4 % or EuroSCORE II < 4 %). In older patients (≥75 years), or those who are inoperable or at high surgical risk (i.e., STS-PROM>8 % or EuroSCORE II > 8 %) TAVR is preferred. All other patients should be assessed for either TAVR or SAVR including other factors: For these patients SAVR is favoured: Lower surgical risk (according to STS-score, EuroSCORE II or logistic EuroSCORE), younger age, transfemoral access challenging or impossible and SAVR feasible, aortic annular dimensions unsuitable for available TAVR devices, bicuspid aortic valve, valve morphology unfavourable for TAVR (e.g. high risk of coronary obstruction due to low coronary ostia or heavy leaflet/LVOT calcification), a thrombus within the left ventricle, significant multi-vessel coronary artery disease requiring surgical revascularization, severe primary mitral valve disease, severe tricuspid disease, significant dilatation/aneurysm of the aortic root and/or ascending aorta and septal hypertrophy requiring myectomy. For these patients TAVR is recommended: higher surgical risk (according to STS-score, EuroSCORE II or logistic EuroSCORE), older age, previous cardiac surgery (particularly intact coronary artery bypass grafts at risk of injury during repeat sternotomy), severe frailty, TAVR feasible via transfemoral approach, sequelae of chest radiation, porcelain aorta, high likelihood of severe patient–prosthesis mismatch (AVA <0.65 cm<sup>2</sup>/m<sup>2</sup> body surface area) and severe chest deformation or scoliosis." |
| summary (Example of a physician-generated case summary) | "The patient is aged 75 years or above*. The logistic EuroSCORE is missing, the EuroSCORE II is 12.99 %, the STS score is missing. The patient does not have coronary heart disease requiring treatment. The patient does not have a left ventricular ejection fraction ≤ 40 %. The patient does not have chronic obstructive pulmonary disease GOLD Stage III or higher. The patient does not have pulmonary arterial hypertension. The patient has active malignancy. The patient does not have liver cirrhosis. The patient does not have a porcelain aorta. The patient does not have significant thorax deformity or scoliosis. The patient does not have a relevant pre-existing comorbidity that, for example, requires immunosuppression. The patient is not status post chest radiation. The patient is not status post cardiac surgery. The patient is not at high risk for prosthesis-patient mismatch. The patient is frail. The patient is not suffering from either cachexia or malignant obesity. " *for data protection reasons, the specific age is not given here |

**Table S7:** LLM completions. Representative LLM responses and their corresponding categorization into a treatment decision. The left column shows the most common responses we retrieved during our experiments.

| Example Completion | Decision |
| --- | --- |
| "Based on the guidelines, the recommended treatment for a patient with severe aortic stenosis would be TAVR (Transcatheter Aortic Valve Replacement)." | TAVR |
| "Based on the guidelines and the patient's characteristics, the recommended treatment would be TAVR (Transcatheter Aortic Valve Replacement)." |  |
| "Based on the guidelines, the recommended treatment for the patient would be SAVR (surgical aortic valve replacement)." | SAVR |
| "The recommended treatment would be SAVR." |  |
| "Based on the given information, the recommended treatment would be SAVR (Surgical Aortic Valve Replacement)." |  |
| "Based on the findings provided, the recommended treatment would be TAVR (Transcatheter Aortic Valve Replacement) or SAVR (Surgical Aortic Valve Replacement)." | Indeterminate |
| "Based on the guidelines, the recommended treatment for a person with severe aortic stenosis would be TAVR (Transcatheter Aortic Valve Replacement) if they are at higher surgical risk or older, and SAVR" |  |
| "Based on the provided information, it is not possible to determine whether TAVR or SAVR would be the recommended treatment without further details." |  |
| "Based on the guidelines provided, the recommended treatment for a person with severe aortic stenosis would be TAVR (Transcatheter Aortic Valve Replacement) if they are older ($\geq 75$ years) or at high surgical risk." | |

**Table S8:** Performance metrics and imputation strategies. To measure concordance and reliability, we used several metrics. We calculated Cohen’s kappa coefficients to quantify agreement. Accuracy, although intuitive, can be highly misleading in classification tasks with a relevant class imbalance as was the case in our study. Intraclass correlation coefficients (ICCs) were used to measure test-retest reliability based on 10 runs per patient. ICCs are undefined in cases of zero variance and can be skewed by a high proportion of indeterminate answers. To address these limitations, we used Shannon entropy as an additional measure to quantify output variation.

| Target | Performance metric | Description | Handling of indeterminate responses | Interpretation |
| --- | --- | --- | --- | --- |
| **Concordance** ("Were the treatment decisions provided by the LLMs concordant with the HT's treatment decisions?") | **Interrater agreement** | Cohen's kappa coefficients were used to measure the interrater agreement between treatment decisions provided by the LLMs and the Heart Team. The LLMs' treatment decisions were aggregated by a majority vote of 10 runs per patient. | Indeterminate responses were classified as "wrong" decisions, meaning they were set to the opposite of the respective HT decision. | Cohen's kappa coefficients $\leq 0$ indicate no agreement, 0.01-0.20 slight, 0.21-0.40 fair, 0.41-0.60 moderate, 0.61-0.80 substantial, and 0.81-1.0 almost perfect agreement.<sup>3</sup> |
|  | **Accuracy** | Accuracy was defined as the proportion of treatment decisions that agreed with the treatment decisions provided by the Heart Team. | Indeterminate responses were classified as "wrong" decisions, meaning they were set to the opposite of the respective HT decision. | Due to the class imbalance in our patient cohort, a model that exclusively outputs the majority class (i.e., "TAVR") achieves an accuracy of 70 %. |
| | **Intraclass correlation coefficient (ICC)** | ICCs were used to measure test-retest-reliability given ten runs per patient.<br>ICCs were calculated based on a one-way random effects, absolute agreement, single-rater model<sup>4</sup> using Python's pingouin package (version 0.5.3). | Indeterminate responses were re-classified as either "TAVR" or "SAVR" through random sampling with replacement and weights corresponding to the prevalence of HT treatment decisions for TAVR and SAVR in our study cohort. | ICCs $< 0.5$ indicate poor, 0.50-0.75 moderate, 0.75-0.90 good, $> 0.90$ excellent test-retest reliability.<sup>4</sup><br>ICCs are undefined if the between cluster variance is zero, e.g., if the LLM output was always "TAVR" for every run and every patient for a particular experiment. |
| | **Entropy** | Shannon entropy<sup>5</sup> was used to assess the output variation within 10 runs per patient as follows<br>$H = -\sum p(k) \log_2 p(k)$,<br>with $k \in ["TAVR", "SAVR", "indeterminate"]$ and reported as mean normalized entropy. | Indeterminate responses were included for the calculation of entropy values since entropy allows to quantify output variation for more than two classes. | Entropy values close to 0 indicate no variation in model output. Entropy values close to 1 indicate maximum output variation. |
| **Bias**<br>("Were the LLMs' treatment decisions biased towards TAVR or SAVR?") | **Frequency bias index (FBI)** | FBI<sup>6</sup> was defined as the ratio of treatment decisions for TAVR given by the LLM and by the Heart Team. | Indeterminate responses were ignored for the calculation of frequency bias indices. | FBI $> 1$ indicates bias towards TAVR, whereas FBI $< 1$ indicates bias towards SAVR. |
| **Decidability** ("How often were the LLMs undecided about treatment?") | **Decidability** | Decidability was quantified as the proportion of determinate to indeterminate treatment decisions. | - | - |

**Table S9:** Performance metrics. Numerical values of the performance metrics portrayed in Figure 2 are shown in addition to the frequencies of accurate, indeterminate and inaccurate treatment recommendations.

| Experiment | Model | Cohen's Kappa coefficient | Frequency Bias Index (FBI) | Intraclass Correlation Coefficient (ICC) | Entropy | Accurate, Indeterminate, Inaccurate |
| --- | --- | --- | --- | --- | --- | --- |
| RAW | BioGPT | -0.18 | 0.95 | 0.73 | 0.32 | (0.32, 0.37, 0.31) |
|  | Mistral | -0.02 | 1.25 | 0.17 | 0.52 | (0.55, 0.2, 0.26) |
|  | Llama 2 | 0.00 | 1.43 | 0.09 | 0.04 | (0.69, 0.01, 0.3) |
|  | PaLM 2 | -0.02 | 1.44 | 1.00 | 0.00 | (0.69, 0.01, 0.3) |
|  | GPT-3.5 | -0.47 | 1.51 | 0.84 | 0.07 | (0.29, 0.54, 0.17) |
|  | GPT-4 | 0.09 | 1.39 | 0.80 | 0.01 | (0.71, 0.02, 0.27) |
|  | GPT-4 Turbo | 0.00 | 1.43 | 0.00 | 0.00 | (0.7, 0.0, 0.3) |
|  | GPT-4o | 0.19 | 1.35 | 0.71 | 0.03 | (0.73, 0.03, 0.24) |
|  | DeepSeek-R1 | 0.22 | 1.30 | 0.19 | 0.17 | (0.75, 0.01, 0.24) |
| RAW+ | BioGPT | -0.31 | 0.90 | 0.81 | 0.19 | (0.29, 0.41, 0.3) |
|  | Mistral | 0.04 | 1.14 | 0.13 | 0.65 | (0.5, 0.24, 0.25) |
|  | Llama 2 | 0.17 | 1.20 | 0.34 | 0.54 | (0.4, 0.5, 0.1) |
|  | PaLM 2 | -0.02 | 1.44 | 1.00 | 0.00 | (0.69, 0.01, 0.3) |
|  | GPT-3.5 | -0.12 | 1.39 | 0.89 | 0.04 | (0.56, 0.24, 0.2) |
|  | GPT-4 | 0.17 | 1.37 | 0.42 | 0.05 | (0.7, 0.03, 0.27) |
|  | GPT-4 Turbo | 0.00 | 1.43 | - | 0.00 | (0.7, 0.0, 0.3) |
|  | GPT-4o | 0.24 | 1.34 | 0.66 | 0.01 | (0.75, 0.02, 0.23) |
|  | DeepSeek-R1 | 0.40 | 1.19 | 0.48 | 0.17 | (0.8, 0.02, 0.18) |
| SUM | BioGPT | 0.04 | 0.82 | 0.27 | 0.46 | (0.29, 0.44, 0.27) |
|  | Mistral | 0.00 | 1.31 | 0.00 | 0.45 | (0.59, 0.12, 0.28) |
|  | Llama 2 | 0.00 | 1.40 | 0.03 | 0.06 | (0.68, 0.0, 0.32) |
|  | PaLM 2 | 0.00 | 1.43 | - | 0.00 | (0.7, 0.0, 0.3) |
|  | GPT-3.5 | -0.25 | 0.90 | 0.85 | 0.12 | (0.41, 0.15, 0.45) |
|  | GPT-4 | 0.44 | 1.15 | 0.69 | 0.12 | (0.81, 0.0, 0.19) |
|  | GPT-4 Turbo | 0.54 | 1.14 | 0.87 | 0.05 | (0.84, 0.0, 0.16) |
|  | GPT-4o | 0.44 | 1.00 | 0.88 | 0.06 | (0.77, 0, 0.23) |
|  | DeepSeek-R1 | 0.34 | 0.66 | 0.44 | 0.34 | (0.65, 0.01, 0.34) |
| SUM+ | BioGPT | -0.02 | 1.14 | 0.15 | 0.61 | (0.55, 0.18, 0.27) |
|  | Mistral | 0.00 | 1.01 | 0.01 | 0.63 | (0.56, 0.07, 0.36) |
|  | Llama 2 | 0.22 | 1.21 | 0.15 | 0.54 | (0.58, 0.26, 0.16) |
|  | PaLM 2 | 0.47 | 1.23 | 1.00 | 0.00 | (0.81, 0.0, 0.19) |
|  | GPT-3.5 | 0.31 | 0.46 | 0.88 | 0.06 | (0.62, 0.0, 0.38) |
|  | GPT-4 | 0.62 | 0.99 | 0.97 | 0.01 | (0.84, 0.0, 0.16) |
|  | GPT-4 Turbo | 0.61 | 1.12 | 0.93 | 0.03 | (0.86, 0.0, 0.14) |
|  | GPT-4o | 0.58 | 1.12 | 0.97 | 0.01 | (0.84, 0, 0.16) |
|  | DeepSeek-R1 | 0.63 | 0.86 | 0.66 | 0.21 | (0.8, 0.01, 0.19) |
| - | Reference Model | 0.55 | 1.11 | 1 | 0 | (0.83, 0, 0.17) |

